# Real-world smartphone data predicts mood after cerebrovascular symptoms and may constitute digital endpoints

**DOI:** 10.1101/2025.01.23.25320624

**Authors:** Stephanie Zawada, Jestrii Acosta, Caden Collins, Oana Dumitrascu, Ehab Harahsheh, Clint Hagen, Ali Ganjizadeh, Elham Mahmoudi, Bradley Erickson, Bart Demaerschalk

## Abstract

Though depression is prevalent in patients with cerebrovascular dysfunction, screening for symptoms is not routine and is often limited via subjective patient-reported surveys. Using smartphone sensors, we sought to evaluate the performance of objective behavior measures and self-report surveys at predicting depression severity in patients with cerebrovascular syndromes. Among enrolled participants (n = 54), 35 patients with ischemic stroke or transient ischemic attack symptoms were monitored in real-world settings using the Beiwe app for 8 or more weeks with adequate compliance. Depression symptoms were tracked via weekly Patient Health Questionnaire-8 (PHQ-8) surveys, monthly personnel-administered Montgomery-Asberg Depression Rating Scale (MADRS) assessments, and passive smartphone sensors. Across weeks, several passive measures were significantly associated with PHQ-8 scores. Personnel-assessed depression severity moderately correlated with self-reported scores. To estimate MADRS, we applied linear mixed models using passive data and PHQ-8 scores. Using antecedent PHQ-8 scores and demographic data, average root-mean-squared error (RMSE) for depression severity prediction across models was 1.54 with accelerometer data, 1.40 also including global position system (GPS) data, and 1.33 also including PHQ-8 open survey duration. Though future research should validate this decentralized approach in a larger cohort, real-world monitoring with active and passive data may triage cerebrovascular patients for efficient depression screening and provide novel mobility and response time outcome measures.

## Introduction

Post-stroke depression (PSD) is well established as a risk factor for mortality, poor functional recovery, and subsequent stroke or cerebrovascular disease (CeVD) diagnoses [1]. Although roughly 1/3 of stroke survivors develop PSD, screening for PSD is not routine. As such, the exact prevalence of PSD remains unknown [2]. While PSD symptoms may initially improve in the first few months, research suggests the majority experience persistent depression in the subsequent years, a period when regular depression screening is not common [3]. Differential diagnosis for PSD is complicated by characteristic post-stroke fatigue as well as the emergence of neurologic deficits, like communication and cognitive difficulties, that can bias self-reported assessments [4]. The traditional classification of stroke as a circulatory system disease has made it challenging to determine whether the stroke-associated depressive symptoms are a predictor or a cause of the condition. Recently, the ICD-11’s reclassification of stroke as a neurological disorder has sparked interest in studying populations with overlapping symptoms, aiming to advance from exploratory research to rigorous scientific investigations with real-world clinical applications [5].

Prior to ICD-11, the majority of research conducted in the intersection of stroke and depression focused exclusively on major stroke cases, with little attention paid to transient ischemic attack (TIA) or neurology admissions for conditions mimicking cerebrovascular ischemia, conditions also linked with depressive symptoms [6, 4]. The wide range of symptoms associated with individual PSD cases as well as non-depressive transient ischemic symptoms challenges the designation of a robust tool for PSD diagnosis and grading. Despite the introduction of several tools for this purpose in the past years, they are not routinely used in practice [7, 8].

Though clinical trials are the key to advancing neurorecovery after stroke, their robust implementation is hindered by insufficient requisite infrastructures, including clinicians, scientists, data analysts, and study staff as well as participant loss to follow-up due to the travel demands for on-site visits [9, 10]. Historically, the identification of PSD has relied upon observation by clinical staff, particularly psychiatrists or neurologists with specialized training in psychiatric conditions in stroke patients; however, a recent meta-analysis found that the standard Patient Health Questionnaire (PHQ-9) is an effective screening tool for PSD, helping triage patients for further diagnostic assessment [11, 12]. Additionally, the PHQ-8, identical to PHQ-9 except for the lack of suicidality assessment, has emerged as a reliable longitudinal measure of PSD symptoms [13].

In recent years, objective and semi-continuous data from real-world monitoring of patients has made strides in understanding cognitive and behavioral changes in lieu of self-reported scores [14]. Powered by low-burden wearable monitors and portable devices, novel data streams of passive sensor and active self-reported measurements are actively being applied to exploratory biomarker research in neurology, assessing psychiatric status and motor function [15]. Research evaluating digital biomarkers to better phenotype and predict disease trajectories with neurological conditions is on the rise, and opportunities exist to validate digital solutions in routine clinical practice [16]. Though the use of wearables to assess post-stroke function has been widely documented [17], few have used continuous remote monitoring in real-world settings to investigate PSD [18–20]. Bui et al. applied an ecological momentary assessment (EMA) delivered via smartphone app 5 times per day over a 2-week period that also required participants (n = 202) to report what activities were undertaken, where, and with whom. In these mild-to-moderate stroke patients, depressed mood was inversely associated with self-ratings of satisfaction, performance, and engagement [21]. Lau et al combined 1-week EMA sampling 8 times per day with accelerometer wear (n = 40), finding that greater depressed affect was associated with increased controlled motivation (β = 0.06, p <.001), caused by external rather than autonomous reasons [22]. Using the Caltrac accelerometer for 6 weeks and in-person record of the depression subscale of the Hospital Anxiety and Depression Scale (HADS), Blaszcz et al. found that, although physical activity increased after hospital discharge, no change in depression scores at baseline and study conclusion was significant (n = 21) [23]. In a cohort of minor stroke patients (n = 76), Ashizawa et al. used the belt-worn Active Style Pro HJA-750C: OMRON accelerometer to measure in-hospital physical activity, and 3 months after discharge, mailed a paper version of Geriatric Depression Scale-15 (GDS-15) to participants. They found direct and inverse relationships between depression and sedentary (OR = 1.130, 95% CI = 1.013 1.281, *p* = 0.028) and light physical activity (OR = 0.853, 95% CI = 0.746 0.976, *p* = 0.021), respectively [24]. Some of our prior work found that real-world monitoring may be more appropriately applied to observing broad changes in cerebrovascular function, rather than exclusively applied to acute events, like stroke [4, 25].

Preliminary research suggests that wearable and portable device monitoring may elucidate phenotypes associated with PSD [26–30]. Despite progress in using wearables to understand behaviors linked with PSD, no clinically actionable system using real-world monitoring has been implemented. Though objective sensor data could help clinicians understand behaviors specific to depression linked with cerebrovascular dysfunction, no research has offered such a digital phenotype, or “moment-by-moment quantification of the individual-level human phenotype in situ” [31]. Among portable devices, smartphone ownership is nearly ubiquitous, with over 6 billion adults owning a smartphone worldwide [32]. Modern smartphones include a set of high-quality sensors, including but not limited to accelerometers, capable of active (e.g., surveys) and passive (e.g., global positioning system or GPS) data collection, positioning them as a potential useful tool for remote biomarker monitoring. Applying smartphones to collect quality longitudinal data appears meaningful for investigating psychiatric and neurologic symptoms linked with chronic disease.

The objective of this study was to evaluate whether a smartphone app could quantify depression after cerebrovascular dysfunction through passive (accelerometer, GPS, and screen touch sensors) and active (surveys) data. In a transdiagnostic cohort of patients with cerebrovascular dysfunction, we aimed to (1) identify digital sensor phenotypes correlated with mood in patients with stroke or TIA symptoms and (2) assess the potential of augmenting current screening processes by using active, passive, and combined active-passive smartphone measures to predict depression severity.

## Methods

### Study Design and Population

This study used a prospective cohort design to recruit participants who were recently admitted to the hospital with ischemic stroke (IS) or TIA symptoms. Participants aged 18 and older who presented at Mayo Clinic Hospital in Phoenix, Arizona were screened for eligibility with the Mayo Clinic Arizona Stroke Monitoring Program using electronic health record (EHR) information and recruited via e-mail after discharge between March and August 2024. We sought to recruit a cohort of transdiagnostic outpatients with similar symptoms at hospital admission and controls with a prior IS or TIA diagnosis participating in sequelae monitoring at Mayo Clinic.

Diagnoses at discharge were confirmed by clinical imaging results based on diagnostic criteria. Participants were included if they had one of the following discharge diagnoses: TIA, IS, and other transient stroke-like symptoms.

Eligible participants were required to own a smartphone with a cellular data service and be able to provide informed consent digitally. Exclusion criteria included those with a prior dementia diagnosis. All participants provided informed consent via digital consent. Mayo Clinic Institutional Review Board approved the study procedure (22-009345).

### Data Collection

Participant registration was conducted remotely. Demographic data was obtained from EHRs. Participants downloaded the HIPAA-compliant digital phenotyping Beiwe app at baseline.

Passive accelerometer, GPS, and screen touch sensor data was recorded along with remote self-entry of the PHQ-8 survey (0-24) using the Beiwe app.

Beiwe is an open-source smartphone platform for digital phenotyping compatible with both iOS and Android operating systems that includes a HIPAA-compliant data storage component and the *Forest* library of data processing packages. The Beiwe app accesses hardware and software within the phone and transmits this raw data to a secure AWS server for standardized processing, handling of data missingness, and imputation. To prevent the linking of personal identifiers and phone data, the Beiwe platform encrypts phone numbers and generates a unique identifier for each participant. The list of passive predictors derived from smartphone sensor streams is available in Table 1.

**Table 1.** Predictors Used in Linear Mixed Models.

| **Baseline Predictors** | **Active Data Predictors** | **Passive Data Predictors** |
| --- | --- | --- |
| Age | Antecedent PHQ-8 Survey Score | GPS (Number of significant locations visited, significant location entropy, time spent at home, distance traveled, maximum diameter of travel, maximum travel distance from home, radius of gyration, average flight length, standard deviation of flight length, average flight duration, standard deviation of flight duration, total time spent in pause/stationary, average stationary/pause time, standard deviation of stationary/pause time, hours per day without GPS data) |
| Sex |  | Accelerometer (Cadence, Walking Time, Number of Steps, hours per day without accelerometer data) |
| Diagnostic Category |  | Touch Screen (Time spent completing antecedent PHQ-8 survey) |
| Initial PHQ-8 score |  |  |
| Prior Depression Diagnosis |  |  |

**Table 2.** Baseline demographic characteristics for participants.

|  | Participants (n = 44) |
| --- | --- |
| **Sex (%)** |  |
| *Male* | 59.1% (26) |
| *Female* | 40.9% (18) |
| **Age (years)** |  |
| *Mean (SD)* | 57 |
| *Min, Q1, Q3, Max* | 23, 49, 68, 83 |
| **Discharge Diagnosis (%)** |  |
| *TIA* | 22.7% (10) |
| *Ischemic Stroke* | 22.7% (10) |
| *Other Transient Stroke-Like Symptoms* | 34.1% (15) |
| *Prior Stroke/TIA Only* | 20.5% (9) |
| **Race (%)** |  |
| *White* | 88.6% (39) |
| *Native American* | 2.3% (1) |
| *Did Not Disclose* | 5.8% (3) |
| *Asian* | 2.3% (1) |
| **Hispanic Ethnicity (%)** | 7.0% (3) |
| **Smartphone Operating System (%)** |  |
| *iOS* | 77.3% (34) |
| *Android* | 22.7% (10) |
| **Baseline PHQ-8 (Mean (SD))** |  |
| *TIA* | 5 (±4) |
| *Ischemic Stroke* | 3 (±3) |
| *Other Transient Stroke-Like Symptoms* | 7 (±6) |
| *Prior Stroke/TIA Only* | 7 (±4) |
| **Heart Disease (%)** | 65.9% (27) |
| **Diabetes (%)** | 24.4% (10) |
| **Multiple Strokes (%)** | 36.6% (15) |
| **Depression (%)** | 26.8% (11) |
| **Anxiety (%)** | 22.0% (9) |

Technical assistance was provided via email and phone, as requested by participants. Data was collected for up to 90 days (and until final checkpoint visit accommodating participant schedules), with some participants opting to provide additional longitudinal data. The app was removed from smartphones upon study completion.

The Beiwe app was configured to send a notification for the PHQ-8 survey to be completed at baseline and once per week throughout the duration of the study. The sampling frequency of unprocessed accelerometer data (3 axes; 10 Hz) was configured to collect in 10 seconds on followed by 10 seconds off cycle. The GPS data collection cycle (1 minute on/10 minutes off) was determined to be optimal for this study of participants from the Mayo Clinic Arizona campus, based on past experience with feasibility study participants who reported feedback on battery drain and smartphone storage issues [32].

Once every 30 days participants completed a virtual interview with study personnel, during which trained personnel administered a Montgomery–Åsberg Depression Rating Scale (MADRS) assessment (0-60). Participants whose smartphone stopped streaming data or who were no longer responsive to the study team were classified as lost to follow-up.

Clinical event data, including hospital admission, death, and documentation of subsequent stroke up to 30 days after final contact with participants (study completion) was extracted from EHRs (Supplemental Table 1).

### Smartphone Sensor Data Processing

All data were downloaded from the AWS server. Raw PHQ-8 survey data was processed individually, converting qualitative answers for individual questions into corresponding numeric scores (0-3) and creating a total PHQ-8 score. Duplicate or empty survey submissions were removed from the local analysis database. Metadata associated with each survey, such as open survey duration time, was calculated and stored in the local analysis database.

Accelerometer data was cleaned using the Oak algorithm, previously validated via 20 public datasets, in the open-source *Forest* Python library. Raw x-, y-, and z-axis data from the accelerometer sensors were converted into gait cadence values with linear interpolation filling gaps in continuous data.

Lastly, the GPS data was processed using the Jasmine algorithm in the *Forest* library. This algorithm applies as validated quality control feature, filtering participants with suboptimal data collected, including those with location coordinates characterized by less than 50 meters of horizontal accuracy [33, 34]. Relying on home location inferred from dataset characteristics, the GPS imputation method implemented via the Jasmine algorithm is published online. Processed GPS data is converted into pause (stationary movement) and flight (straight-line movement) events, with average, total, and standard deviation metrics calculated for both on a minute-by-minute basis. Information about each GPS measure derived is available in Supplementary Table 2.

To adjust for smartphone sensor data missingness, we calculated the sum of daily minutes the GPS was turned off. Reasons for GPS data missingness are that participants may turn off their phones or disable GPS location streaming, forget to charge their phones, install updates, or use apps conflicting with Beiwe data collection. Additionally, sampling rates may automatically decrease when phones are in a low battery state. For the accelerometer sensor, we also calculated the minutes of missing data per day. A summary table of data completeness by sensor for each participant is in Supplementary Table 3.

### Statistical Analyses

Baseline characteristics along with study compliance metrics were obtained.

To identify digital sensor phenotypes correlated with mood in patients with stroke/TIA symptoms (aim 1), we conducted a repeated measures correlation analysis between sensor predictors and PHQ-8 scores. For each PHQ-8 survey completed, the weekly average of each passive predictor was computed for the 7 days prior to a survey. Using the *rmcorr* package in R, repeated measures correlation generates the within-subject association between two variables using analysis of covariance, a useful tool for longitudinal data.

To assess the use of passive smartphone measures in real-world settings as proxies for depression severity scales typically used in clinical settings (aim 2), linear mixed models (LMMs) were used to predict MADRS scores. LMMs are uniquely able to handle data collected at different timepoints from the same subjects, also known as clustered data, with varying quantities of missing data. Using a modified approach to Pellegrini et al.’s digital phenotyping study focused on psychiatry patients with major depressive disorder, bipolar disorder, and schizophrenia or schizoaffective disorder, we developed 6 models for predicting staff-administered scores (MADRS) [28]. The initial PHQ-8 score of the study was used as a baseline predictor to account for the modest sample size of this pilot study and to simulate realistic conditions at hospital discharge without staff-administered MADRS scores. Considering that the passive predictors were correlated with one another, principal component analysis (PCA) was used to obtain a principal component (PC1) predictor from weekly averages across predictors. The missingness variables for accelerometer and GPS sensors described above were included in PCA.

Before fitting the model, MADRS assessment instances were assessed for completeness, meaning that any instance with missing predictor values was excluded from analysis. Leave-one-subject-out cross validation was performed for each participant, with the model fitted with data from other participants. Then, MADRS scores for the excluded participant were predicted with the model. Model 1 used only the antecedent PHQ-8 score to predict MADRS score; Model 2 used only passive data (weekly average PC1); Model 3 used both the PHQ-8 score and passive data PC1; Model 4 applied the predictors from Model 3 plus the open survey time (screen sensor) predictor; Model 5 used no predictors; and Model 6 included the sensor measure with the highest loading in PC1.

To assess the fit of each model, the root-mean-squared error (RMSE) for each participant was calculated by taking the square of the error between the predicted MADRS and actual MADRS scores for each MADRS instance.

### Sensitivity Analysis

First, we compared smartphone PHQ-8 scores with staff-administered MADRS scores. For each MADRS score, the antecedent PHQ-8 score was utilized for comparison. Given that both PHQ-8 and MADRS scores are ordinal, Spearman’s rank order correlation coefficient was used to obtain regression coefficients and p-values across the 3 MADRS timepoints.

For the repeat measures correlations of Aim 1, we completed a sensitivity analysis investigating correlates between behavior components of PHQ-8 (disinterest, depression, sleep, lethargy, self-blame, appetite, concentration, movement/speech) and smartphone sensor measures.

Taking into account that accelerometer sensors are less data-and battery-intensive than GPS, we repeated the model development process for aim 2 applied to accelerometer-only passive data, including the principal component analysis step. The results of such an investigation have implications beyond smartphone-based digital phenotyping, considering that the evolving suite of wearable wristwatches include built-in accelerometer sensors. Moreover, no longitudinal study of this duration has been applied to mood monitoring for cerebrovascular dysfunction, with the closest comparative study design involving older adult patients with varying levels of cognitive impairment [35].

## Results

### Participant Characteristics

Of the 54 participants enrolled (Figure 1), 4 were unresponsive after completing the digital consent form, 2 dropped out due to medical emergencies/hospitalization, 1 reported data privacy concerns, and 3 dropped out for smartphone-related issues. Among participants who downloaded the app (81.5%), 5 were lost to follow-up due to unresponsiveness prior to the initial MADRS scoring and 1 requested to leave the study. Of the 38 participants who completed the initial MADRS scoring (Figure 1), 3 dropped out: 1 due to chemotherapy, 1 due to subsequent hospitalization, and 1 due to a smartphone issue. Participants who completed the study had MADRS scores recorded on a monthly basis, around the 30-day and 60-day marks, as well as around the 90-day mark if still participating, accommodating participants’ schedules. Of the 44 participants contributing data for the sensor phenotyping analysis (aim 1), 35 were included in prediction models (aim 2).

**Figure 1.**
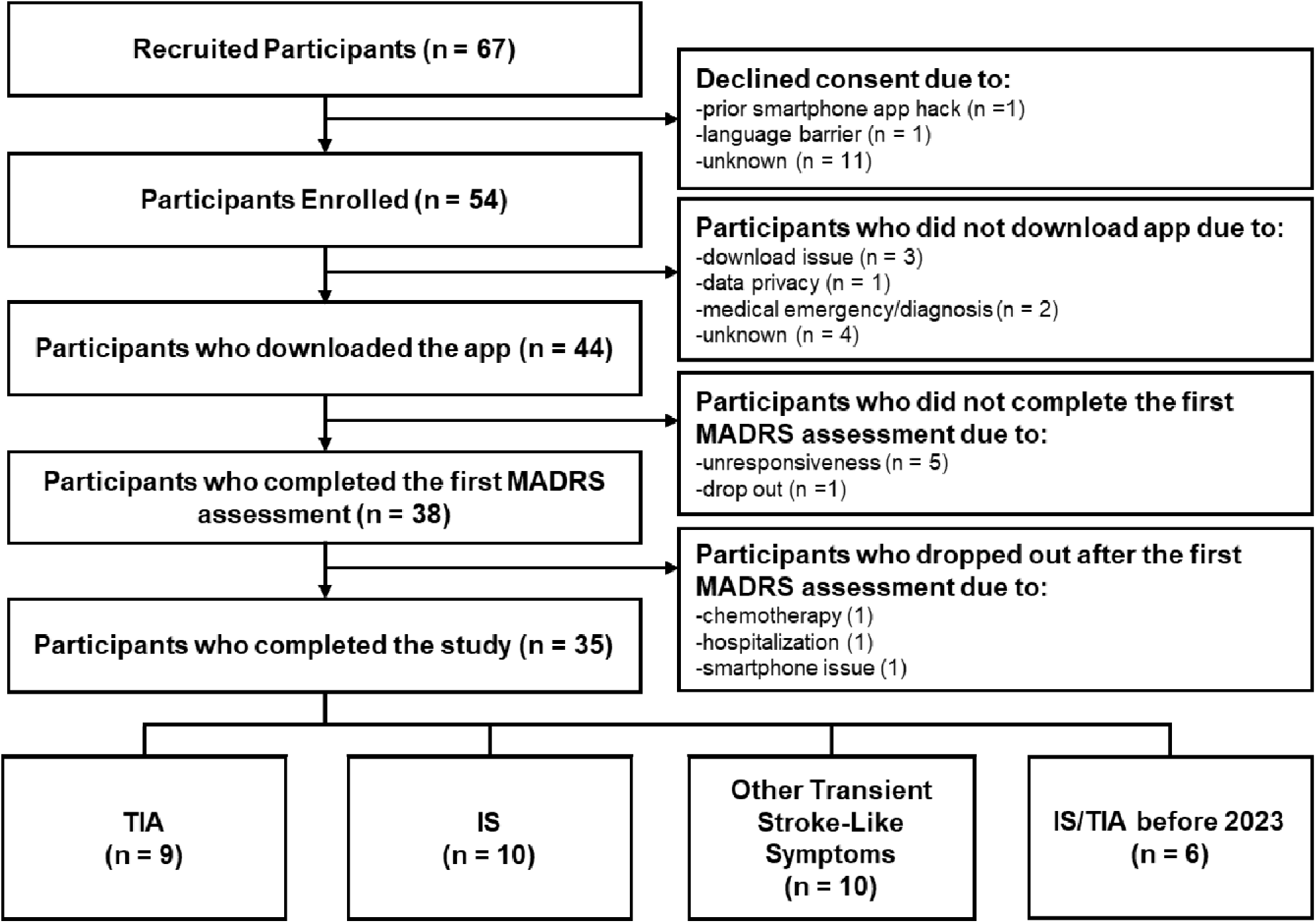
Diagram of study enrollment and retention

Study participants were primarily male (59.1%), middle-aged (57 y), and White (88.6%). Only 7.0% of participants reported Hispanic ethnicity. Most participants had heart disease (65.9%) and 36.6% had suffered multiple strokes/TIAs. At baseline, 24.4% were diabetic, 22.0% had an anxiety diagnosis, and 26.8% had a depression diagnosis.

Most participants had an iOS smartphone (77.3%). Participants in the other transient stroke-like symptoms category at hospital discharge and those with a prior stroke/TIA had an average PHQ-8 score indicating moderate depression (6-8) while participants with an IS or TIA diagnosis at discharge had an average score suggesting mild depression (3-5).

During the study, 6 participants reported surveys indicating major depression and 10 indicated minor depression. Relevant clinical outcomes, including hospitalization for stroke-related conditions or death, were tracked for 30 days after smartphone study completion. Only 2 participants in the study presented at the emergency room for stroke-related symptoms during the study, both of whom completed 90 days of smartphone monitoring (Supplementary Table 1).

Survey completion rates over time were moderately high, with 73% of participants completing 8 or more weeks of PHQ-8 surveys (Supplementary Figure 1); however, PHQ-8 trajectories were highly variable over the study period (Supplementary Figure 2).

### Identification of Digital Sensor Phenotypes Correlated with Mood in Patients with Stroke Symptoms

We examined correlations between mood behaviors measured in the PHQ-8 survey and sensors using repeated measures (n =44). All sensor predictors were the average of 7 days prior to the weekly PHQ-8 survey. Assessing relationships between smartphone accelerometer sensor predictors and PHQ-8 scores (Table 3), PHQ-8 total score was positively correlated with Time to Complete Survey (*r* = 0.151; *p-*value = 0.004) and Distance from Home (*r* = 0.173; *p-*value = 0.008) and negatively correlated with Home Duration (*r =*-0.147; *p*-value = 0.024).

**Table 3.** Repeat Measures Correlates of PHQ-8 Score and Smartphone Measures.

| Smartphone Sensor Measure | Correlation coefficient (r) | *p* | 95% CI |
| --- | --- | --- | --- |
| Time to Complete Survey | **0.151** | **0.004\*\*** | **(0.048, 0.251)** |
| Distance Diameter | 0.013 | 0.848 | (-0.115, 0.140) |
| Distance from Home | **0.173** | **0.008\*\*** | **(0.057, 0.294)** |
| Distance Traveled | 0.014 | 0.831 | (-0.114, 0.141) |
| Flight Distance Average | 0.008 | 0.901 | (-0.119, 0.135) |
| Flight Distance StdDev | 0.021 | 0.745 | (-0.106, 0.148) |
| Flight Duration Average | -0.066 | 0.312 | (-0.192, 0.062) |
| Flight Duration StdDev | -0.094 | 0.148 | (-0.219, 0.034) |
| Home Duration | **-0.147** | **0.024\*** | **(-0.269, -0.020)** |
| Gyration Radius | 0.006 | 0.923 | (-0.121, 0.134) |
| Significant Location Count | -0.055 | 0.400 | (-0.181, 0.073) |
| Significant Location Entropy | -0.070 | 0.283 | (-0.196, 0.058) |
| Pause Time | -0.081 | 0.212 | (-0.207, 0.047) |
| Total Flight Time | -0.054 | 0.410 | (-0.180, 0.074) |
| Pause Duration Average | -0.014 | 0.826 | (-0.142, 0.113) |
| Pause Duration StdDev | 0.025 | 0.704 | (-0.103, 0.152) |
| Walking Time | -0.026 | 0.645 | (-0.137, 0.085) |
| Steps | -0.028 | 0.623 | (-0.139, 0.083) |
| Cadence | -0.024 | 0.669 | (-0.135, 0.087) |

### Assessment of passive smartphone measures as proxies for post-stroke depression severity

Before building models with passive accelerometer and GPS data, one participant was excluded due to sampling below the quality check threshold for the *Forest* package. For the remaining participants (n = 34), principal component analysis was performed to obtain the first principal component (PC1) (Supplementary Figure 3). In these models, PC1 was used as a predictor. The components of PC1 and their respective loadings are listed in Supplementary Table 4. PC1 was responsible for 40.9% of data variance, and GPS data contributed PC1’s highest loading measures.

In the models assessing the potential of smartphones measures to predict MADRS scores (Table 4), the lowest RMSE was 1.33 (Model 4 with demographics), which included accelerometer, GPS, screen sensor data and the antecedent self-report PHQ-8 score.

**Table 4.** RMSE predicting MADRS scores using Models 1-6 with three variations (no demographics; with demographics; with demographics + initial PHQ-8 scores)

|  |  | **+demographics** | **+demographics**<br>**+initial PHQ-8** |
| --- | --- | --- | --- |
| **Model 1 (PHQ-8 Score)** | 1.67 | 1.54 | 1.56 |
| **Model 2 (Accelerometer + GPS)** | 1.68 | 1.52 | 1.54 |
| **Model 3 (Accelerometer + GPS + PHQ-8 Score)** | 1.59 | 1.40 | 1.46 |
| **Model 4 (Accelerometer + GPS + PHQ-8 Score + Survey Timing)** | 1.57 | 1.33 | 1.46 |
| **Model 5 (Only Participant Effect)** | 1.69 | 1.58 | 1.64 |
| **Model 6 (Significant Location Count)** | 1.73 | 1.55 | 1.58 |

In the model variation without demographic variables, the average RMSE for Model 1 was 1.67; 1.68 for Model 2; 1.59 for Model 3; 1.57 for Model 4; 1.69 for Model 5; and 1.73 for Model 6. Here, the inclusion of passive smartphone predictors did improve the prediction of MADRS scores compared to using antecedent PHQ-8 self-report scores only. This trend was observed across model variations, including the variation with demographic variables and the variation with demographic variables and initial PHQ-8 scores. The model variation with the best performance included those with the demographic baseline variables; including the initial PHQ-8 score, simulating discharge screening after hospitalization, in the model variation with demographic variables did not improve model fit.

In the variation with demographic data only, the inclusion of passive smartphone data with the antecedent PHQ-8 score yielded a modest RMSE improvement of 0.12 compared to the model with only passive data; however, including the PHQ-8 score survey timing (seconds) improved the fit of this passive-active data model prediction by 0.07.

Model 6 assessed the measure that contributed the highest loading in PC1 without including the PC1 itself; however, isolating and including this measure alone did not improve model fit relative to the passive-active models.

### Sensitivity Analysis

In aggregate, antecedent PHQ-8 scores moderately correlated with personnel-administered MADRS scores (*r* = 0.667), a finding supported by prior literature (Supplementary Figure 4). The correlations between antecedent PHQ-8 scores and MADRS scores, across assessments were as follows: 0.599 at initial assessment (Supplementary Figure 5a), 0.700 at the second assessment (Supplementary Figure 5b), and 0.75 at the third assessment (Supplementary Figure 5c).

Regarding Aim 1, the repeat measures correlations between smartphone measures and individual components of the PHQ-8 survey revealed the following: lethargy was negatively correlated with significant location count (*r =*-0.162; p-value = 0.013) and significant location entropy (*r =* - 0.133; p-value = 0.040); appetite changes were positively correlated with the (max) distance from home (*r =* 0.275; p-value < 0.001) and negatively correlated with time spent at home (*r =* - 0.274; p-value < 0.001); trouble concentrating was positively correlated with (max) distance from home (*r =* 0.141; p-value = 0.031); and changes in speech/movement was positively correlated with (max) distance from home (*r =* 0.306; p-value < 0.001) and negatively correlated with time spent at home (*r =*-0.285; p-value < 0.001) and total stationary/pause time (*r =* - 0.172; p-value = 0.008).

For the accelerometer-only MADRS prediction models, the model with accelerometer data and antecedent PHQ-8 scores performed worse than any of the GPS-accelerometer models in Table 4. The model including only demographic data and the antecedent PHQ-8 score (RMSE = 1.51) outperformed the accelerometer-only models.

## Discussion

Our exploration of digital phenotyping using the Beiwe application to understand real-world behavior of patients with cerebrovascular syndromes contributes to scientific rationales for remote monitoring of post-stroke/TIA patients.

This pilot study offers initial feasibility data for smartphone-based digital phenotyping and monitoring in patients with cerebrovascular syndromes. We demonstrated that longitudinal measures derived from smartphone sensors may serve as proxies for mood in patients with cerebrovascular symptoms, a finding with implications for PSD screening, clinical outcome assessments, and digital endpoints in decentralized clinical trials. Overall, our findings complement previous cerebrovascular disease studies using ecological momentary assessment (EMA), namely that adherence to smartphone-based studies is high in this population regardless of depression severity, age or post-diagnosis functional ability [36]. Additionally, these results extend those that found a close association between self-reported mood and functional ability in post-stroke patients [37].

While digital phenotyping has been applied to investigating depression and anxiety in a growing number of clinical populations, ranging from healthy older adults to psychiatric patients with severe symptoms}, this is the first study to investigate its application to IS, TIA or cerebrovascular syndromes, to our knowledge [30]. Considering that cerebrovascular syndromes are heterogenous by nature, the need for technologies to advance personalized care is critical, and real-world monitoring with passive and active data, like the approach presented in this study, may offer a path forward. Specifically, we found mild correlations between some smartphone measures and PHQ-8. Screen-based sensors that capture time spent completing the PHQ-8 survey may augment screening for depression, considering the mild positive correlation between time-to-survey-completion and PHQ-8 score. While psychomotor impairment is well documented in depressed populations, digitally-based time-to-decision-making measures, like open survey completion/read times, have been shown to classify participants with cognitive impairment with higher accuracy than traditional assessments [38, 39]. The negative correlation between time spent at home and PHQ-8 score could be characteristic of post-stroke/TIA patients. Considering the majority of our study’s participants were admitted to the hospital for stroke/TIA symptoms within the year of enrolling in the study, it is expected this population would be spending more time at home than others [40]. Simultaneously, the positive correlation between (max) distance from home and both PHQ-8 score and trouble concentrating may reflect the burden of longer travel or commute demands on patients who have suffered a stroke/TIA or similar symptoms.

Traveling to fewer locations (significant location count) and spending less time at those locations (significant location entropy) is congruent with increased feelings of lethargy, also characteristic of post-stroke fatigue (PSF). Combined with the correlates with overall PHQ-8 scores, this smartphone-based phenotype may shed new light on PSF, a condition for which few measurement tools have been validated and limited evidence for successful interventions exists [41]. Considering that the question assessing movement and speech asks about the frequency of change, rather than its direction, the correlations with pause time, max distance from home, and time spent at home are congruent with the question’s probing line: “moving around a lot more than usual”. These findings suggest GPS measures of mood in post-stroke/TIA patients could have application in rehabilitation and physical therapy contexts, offering personalized insights into PSF. Further support for this hypothesis that digital phenotyping in cerebrovascular dysfunction may capture PSF are findings from Hackett et al.’s study of older adults (including those with and without cognitive impairment) using the mindLAMP smartphone app (n = 37). In contrast to our findings, Hackett et al. found that more location diversity was associated with less depression, in this comparative sample of patients without a recent stroke [35].

While the inclusion of accelerometer passive measures with PHQ-8 scores did not improve predictive performance, the use of accelerometer measures in conjunction with GPS and screen sensor measures did. One reason for this small improvement could be that cerebrovascular syndrome patients experience slight behavior aberrations due to cognitive changes that go unnoticed. This finding contributes clinically meaningful information for future study designs involving smartphones, highlighting the potential for multimodal sensor data to offer a more comprehensive observation of patients. For example, while passive accelerometer data alone may not help clinicians identify patients at risk of depression, the combination of multiple sensors may offer new insights that transform remote monitoring into clinically actionable insights. The improved prediction performance of passive data is also supported by Lau et al’s one-week wearable accelerometer study with mild stroke patients and Yi et al’s 12-month Beiwe study with nurses (n = 2394), finding that self-reported mood and activities correlate strongly with accelerometer measures [21, 42]. A key strength of this study is that it demonstrates the potential for smartphone accelerometer sensors to collect longitudinal data. One critical issue with wearable accelerometers is the participant burden of wearing and charging the device over sustained periods of time, and our results suggest smartphone monitoring could fill in gaps in data collection for wearables or be offered as an alternative to participants in future studies.

Additionally, these findings extend those of Pellegrini et al., which evaluated the performance GPS and accelerometer data in predicting MADRS scores in a transdiagnostic population of psychiatric patients. In contrast to Pellegrini et al, we found that passive (GPS and accelerometer) data did improve MADRS score prediction; however, the range of PHQ-8 and MADRS scores in our cohort was smaller than theirs, as reflected in the lower RMSE values for our models. Moreover, we exclusively examined smartphone-based accelerometer data in our sensitivity analysis and found that accelerometer data, which is less draining on smartphone batteries and more frequently sampled, did not improve MADRS prediction over using antecedent self-report PHQ-8 scores alone. Unlike the cohort of psychiatric patients, our cohort of patients with cerebrovascular syndromes have exhibited a range of functional changes, from numbness to one-side loss of function. The results of our cohort outline a potentially meaningful use case for using multiple smartphone sensors to monitor behavior associated with brain changes and could open new avenues of research at the intersection of computational neuroimaging and real-world validity [43, 44]. Future research should compare the effectiveness of wearable sensor combinations at augmenting depression and fatigue screening for patients with cerebrovascular disease symptoms.

This work is subject to numerous limitations and should be considered as a starting point for future work. First, the pilot study design includes a small sample size when considering effect, although the sample size is average for smartphone studies. Second, passive measures in this analysis were not processed to consider weekday versus weekend averages. Next, participants most likely did not carry their phones with them constantly. As such, differences in participant data could be partly driven by phone use patterns. Though the use of time-averaged measures is standard in smartphone research, little is known about optimal timepoints for analysis, i.e. using an average from 3 days before an assessment versus the average of 7 days prior [30]. Moreover, the interpretation of GPS data should be considered carefully, because of the on-off cycles required for its successful collection in real-world settings when patients need reliable access to their phones. The behaviors assessed via self-report are limited by the PHQ-8. Originally, we planned to use the Beck Depression Inventory (BDI-II) measurement for weekly survey collection; however, our feasibility study found that the BDI-II was too burdensome for post-stroke/TIA patients. Another challenge documented was battery drain associated with GPS streaming and routine OS software updates; yet, emerging research suggests battery drain may also be a digital biomarker of cognitive function [45]. Some participants also required frequent password and technical assistance, a factor that future researchers could quantify in cognitive digital health studies. Multiple participants traveled for vacations and work, a real-world consideration that further complicates data collection. Missing data due to missed surveys, battery drain, and sick days occurred, but are common in decentralized research. Additionally, the results of the study may not generalize well to non-White patients. As most participants had an iOS smartphone, data collection may also vary in ways unnoticed. Beyond issues associated with wireless and cellular access in rural areas in Arizona, participant travel for vacation or work to different cities may also have introduced unforeseen variations in data. Though not a limitation, we chose to take the average of 7 days for each measure in our analyses, as is commonly found in the growing set of studies in this space; however, it is plausible that other time periods not explored in this study may be more useful at predicting scores [30]. Lastly, it is important to consider that the improved model fit offered by the inclusion of passive measures, compared to self-reported scores alone, is small (RMSE decrease of 0.21) and may be more useful if applied to predicting scores during weeks of lapsed survey compliance rather than for screening purposes. A larger sample population is necessary to investigate the optimal role of remote monitoring with smartphones in this context. Since the model variations including the initial PHQ-8 scores did not improve score prediction, more recent data samples appear to be more useful than baseline self-report scores, offering further support for mood monitoring of cerebrovascular syndromes in real-world settings.

Overall, this study presents a novel and more robust look at real-world mood in patients with recent IS, TIA or stroke-like symptoms. The choice to include patients with stroke-like symptoms highlights the usefulness of such a monitoring program in a neurology clinic beyond stroke care. This study also demonstrates the potential of frequent and low-burden sampling of mood via surveys without visiting a clinical site, an important consideration for patients with motor impairment or those who reside in rural areas. Additionally, the collection of objective captures of physical function holds promise for better understanding individual patient outcome trajectories.

## Conclusions

Our study suggests that smartphone GPS measures may serve as proxies for mood and elucidate new phenotypes of mood in patients with cerebrovascular syndromes. Passive GPS, accelerometer, and screen sensor data slightly improved the prediction of medical personnel-graded mood scores compared to patient-reported surveys alone, outlining a potentially meaningful use case for passive behavior monitoring in patients with cerebrovascular syndromes to augment screening protocols. Our preliminary work lays a foundation for using mobility, spatial, and mobile device screen data to investigate environmental and behavioral factors influencing mood in patients with cerebrovascular syndromes. Future research should involve larger cohorts to identify which measures are most meaningful for mood prediction, to compare the effectiveness of different combinations of wearable sensors, and to establish clinically useful thresholds for triaging patients at different timepoints in recovery.

## Data Availability

All data produced in the present study are available upon reasonable request to the authors

