## Supplementary Materials for "Real-world smartphone data predicts mood after cerebrovascular symptoms and may constitute digital endpoints"

Supplementary Table 1. Clinical outcomes over the 90-day study completion.

| **Outcome** | **Completed 30 days (n = 3)** | **Completed 60 days (n = 38)** | **Completed 90 days (n = 38)** | **Total (n = 41)** |
| --- | --- | --- | --- | --- |
| **90-day Death, n (%)** |  |  |  |  |
| *Yes* | 0 | 0 | 0 | 0 |
| *No* | 100% (3) | 100% (38) | 100% (38) | 100% (38) |
| *Unknown* | 0 | 0 | 0 | 0 |
| **90-day ED admission or hospitalization for stroke-related condition, n (%)** |  |  |  |  |
| *Yes* | 0 | 0 | 5.3% (2) | 5.3% (2) |
| *No* | 100% (3) | 100% (3) | 94.7% (36) | 94.7% (36) |
| *Unknown* | 0 | 0 | 0 | 0 |

Supplementary Table 2. Statistical Summaries (Daily) Generated by Forest Package (table copied from https://forest.beiwe.org/en/latest/ )

| **Variable** | **Type** | **Description of Variable** |
| --- | --- | --- |
| date | str | Time of observation (_gait_daily.csv format: yyyy-mm-dd; _gait_hourly.csv format: yyyy-mm-dd HH:MM:SS’) |
| walking_time | int | Total walking time (in seconds) |
| steps | int | Total steps taken |
| cadence | float | Average cadence in time window (daily or hourly) |
| Observed duration | Float | The total time when the GPS is on (in hours for daily and minutes for hourly) |
| Observed duration in day | Float | The total time when the GPS is on from 8AM to 8PM (in hours for daily) |
| Observed duration at night | Float | The total time when the GPS is on from 8PM to 8AM (in hours for daily) |
| Home time | Float | Time spent at home over the course of a day (in hours for daily and minutes for hourly) |
| Distance traveled | Float | Total distance travelled over the course of a day (in km) |
| Radius of gyration | Float | Average radius that a person travels from their center over the course of a day (in km) |
| Maximum diameter | Float | Largest distance between any two places that a person visited in a day (in km) |
| Maximum distance from home | Float | Largest distance between any places that a person visited in a day and their home (in km) |
| Number of significant locations | int | Number of significant visited at any point over the course of a day |
| Total flight time | Float | Total time spent in flight over the course of a day (in hours) |
| Average flight length | Float | Average of the length of all flights (straight line movement) that took place over the course of a day (in km) |
| Standard deviation of flight length | Float | Standard deviation of the length of all flights (straight line movement) that took place over the course of a day (in km) |
| Average flight duration | Float | Average of the duration of all flights (straight line movement) that took place over the course of a day (in hours for daily and minutes for hourly) |
| Standard deviation of flight duration | Float | Standard deviation of the duration of all flights (straight line movement) that took place over the course of a day (in hours for daily and minutes for hourly) |
| Total pause time | Float | Total time spent in pause over the course of a day (in hours for daily and minutes for hourly) |
| Average pause duration | Float | Average of the duration of all pauses that took place over the course of a day (in hours for daily and minutes for hourly) |
| Standard deviation of pause duration | Float | Standard deviation of the duration of all pauses that took place over the course of a day (in hours for daily and minutes for hourly) |
| Significant location entropy | Float | Entropy measure based on the proportion of time spent at significant locations over the course of a day |

Supplementary Table 3. Data Missingness for Sensors by Participant

| **Participant No.** | **GPS completeness (Daily)** | **Accelerometer completeness (Daily)** |
| --- | --- | --- |
| 1 | 4.99% | 49.90% |
| 2 | 6.27% | 15.50% |
| 3 | na | 33.90% |
| 4 | N/A | na |
| 5 | NA | 27.00% |
| 6 | 42.4 | 43.00% |
| 7 | 5.25% | 65.10% |
| 8 | 14.6 | 71.00% |
| 9 | 6.4 | 45.90% |
| 10 | NA | 43.30% |
| 11 | 5.16 | 41.20% |
| 12 | NA | 45.20% |
| 13 | NA | 40.20% |
| 14 | 6.37 | 50.30% |
| 15 | 3.66 | 47.00% |
| 16 | 5.45 | 46.50% |
| 17 | 17 | 46.50% |
| 18 | 4.39 | 58.40% |
| 19 | NA | 46.10% |
| 20 | 15.5 | 55.40% |
| 21 | 4.7 | 54.20% |
| 22 | 4.74 | 58.30% |
| 23 | 4.48 | 55.10% |
| 24 | NA | NA |
| 25 | 4.05 | 46.90% |
| 26 | 4.97 | 63.30% |
| 27 | 4.2 | 45.40% |
| 28 | 4.53 | 94.30% |
| 29 | 5.68 | 37.20% |
| 30 | 3.4 | 48.60% |
| 31 | 4.52 | 51.70% |
| 32 | 4.09 | 45.80% |
| 33 | 3.12 | 40.90% |
| 34 | 3.78 | 13.30% |
| 35 | 5.68 | 40.30% |
| 36 | 4.17 | 37.90% |
| 37 | NA | 40.00% |
| 38 | 3.86 | 47.90% |
| 39 | 5.3 | 53.90% |
| 40 | 5.18 | 63.30% |
| 41 | 4.8 | 49.90% |
| 42 | 2.9 | 16.90% |
| 43 | 4.71 | 53.10% |

Supplementary Figure 1. Number of Weeks with Participants who Completed Surveys

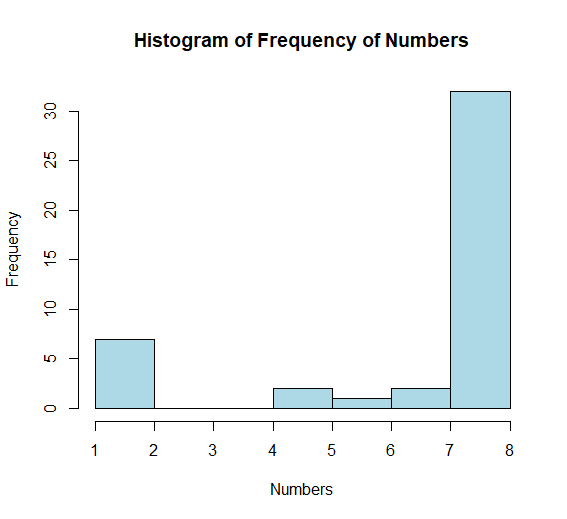

Supplementary Figure 2. Individual PHQ-8 Score Trajectories by Week.

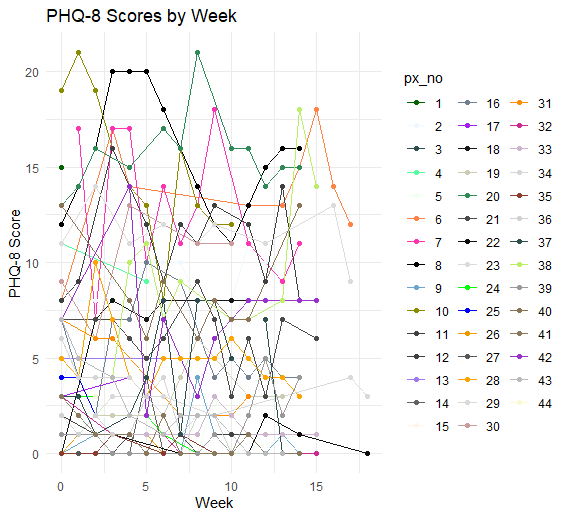

Supplementary Figure 3. PCA Loadings

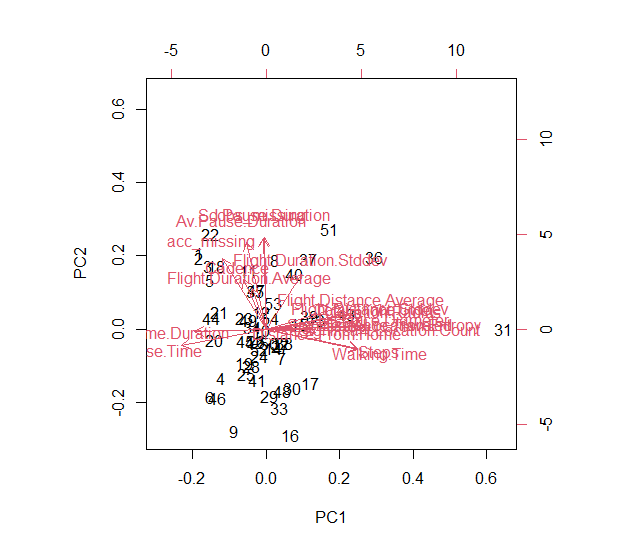

Supplementary Table 4. Principal component analysis loadings

|  | PC1 |
| --- | --- |
| Distance.Diameter | 0.2956573 |
| Distance.From.Home | 0.1541413 |
| Distance.Traveled | 0.3085011 |
| Flight.Distance.Average | 0.2382564 |
| Flight.Distance.Stddev | 0.2613729 |
| Flight.Duration.Average | -0.0429423 |
| Flight.Duration.Stddev | 0.1095711 |
| Home.Duration | -0.2226121 |
| Gyration.Radius | 0.3007212 |
| Significant.Location.Count | 0.3091059 |
| Significant.Location.Entropy | 0.2949143 |
| Pause.Time | -0.2650158 |
| GPS.Missing | -0.0084687 |
| Total.Flight.Time | 0.268787 |
| Av.Pause.Duration | -0.0607645 |
| Sd.Pause.Duration | -0.005339 |
| Walking.Time | 0.284716 |
| Steps | 0.2845166 |
| Cadence | -0.0709859 |
| Acc.Missing | -0.1374335 |

Supplementary Figure 4. Plot for Aggregate PHQ-8 and MADRS Correlation

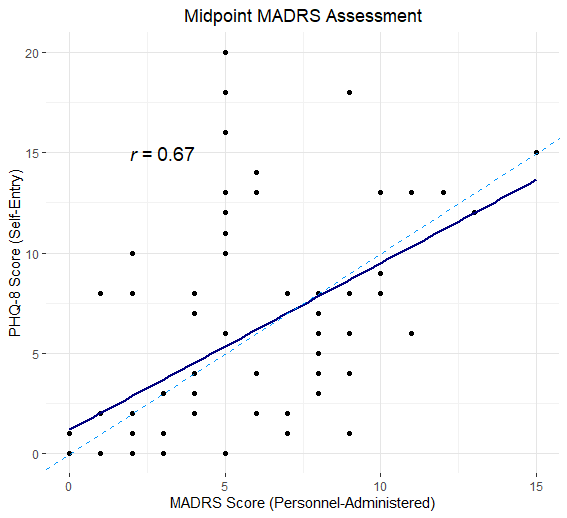

Supplementary Figure 5a. Plot for PHQ-8 and MADRS Correlation at Timepoint 1

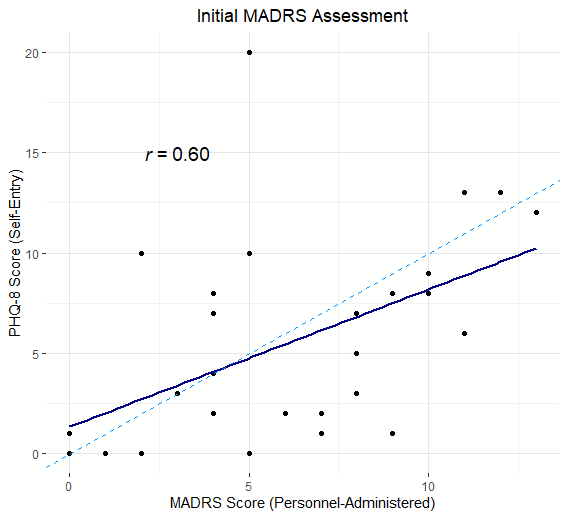

Supplementary Figure 5b. Plot for PHQ-8 and MADRS Correlation at Timepoint 2

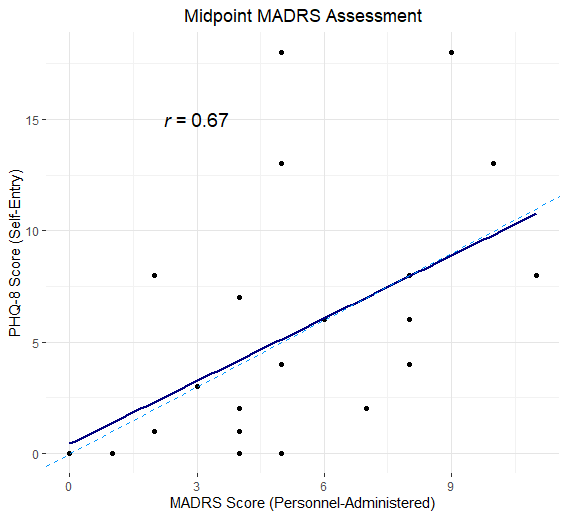

Supplementary Figure 5c. Plot for PHQ-8 and MADRS Correlation at Timepoint 3

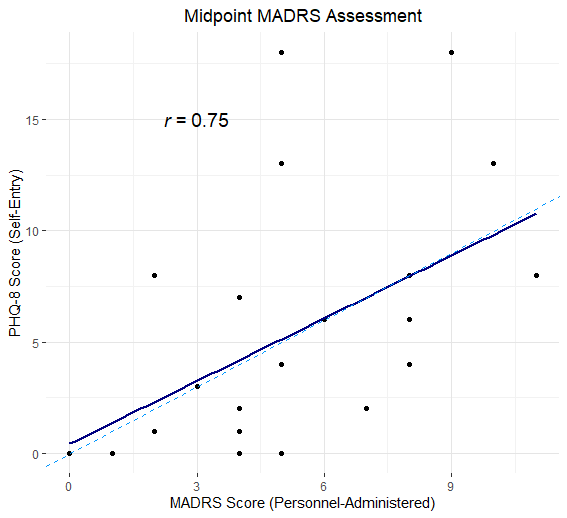

Supplementary Table 5. RMSE predicting MADRS scores using Models 1-6 with three variations (no demographics; with demographics; with demographics + initial PHQ-8 scores)

|  |  | **+demographics** | **+demographics**  **+initial PHQ-8** |
| --- | --- | --- | --- |
| **Model 1 (PHQ-8 score)** | 1.66 | 1.51 | 1.53 |
| **Model 2 (accelerometer data)** | 1.77 | 1.63 | 1.64 |
| **Model 3 (accelerometer + PHQ-8 score)** | 1.70 | 1.54 | 1.57 |
| **Model 4 (accelerometer + PHQ-8 score + survey timing)** | 1.69 | 1.57 | 1.57 |
| **Model 5 (only participant effect)** | 1.74 | 1.59 | 1.60 |
| **Model 6 (Significant.Location.Count )** | 1.77 | 1.62 | 1.62 |
